# Patient factors influencing participation in aerobic exercise during inpatient and outpatient rehabilitation post stroke: a prospective cohort study

**DOI:** 10.64898/2026.08.26.26361451

**Authors:** Azadeh Barzideh, Augustine J. Devasahayam, Susan Marzolini, Sarah Munce, Kathryn M. Sibley, Elizabeth L. Inness, Avril Mansfield

**Affiliations:** Rehabilitation Sciences Institute, University of Toronto, Toronto, ON, Canada; KITE-Toronto Rehabilitation Institute, University Health Network, Toronto, ON, Canada; Evaluative Clinical Science, Hurvitz Brain Sciences Program, Sunnybrook Research Institute, Toronto, ON, Canada; Department of Physical Therapy, University of Toronto, Toronto, ON, Canada; Faculty of Kinesiology and Physical Education, University of Toronto, Toronto, ON, Canada; Holland Bloorview Kids Rehabilitation Hospital, Bloorview Research Institute, Toronto, ON, Canada; Department of Community Health Sciences, Rady Faculty of Health Sciences, University of Manitoba, Winnipeg, MB, Canada; George and Fay Yee Centre for Healthcare Innovation, Winnipeg, MB, Canada

**Keywords:** Stroke, Aerobic exercise, Self-efficacy, Depressive symptoms, Apathy, Rehabilitation, Patient preference

## Abstract

**Background:** Aerobic exercise is recommended during stroke rehabilitation to improve cardiorespiratory fitness and support recovery; however, participation rates remain low. While institutional and system-level barriers have been widely examined, less is known about how individual patient factors influence engagement in aerobic exercise during rehabilitation.

**Objectives:** We aimed to determine whether depressive symptoms, apathy, self□efficacy and outcome expectations for exercise, perceived barriers, or past exercise history were associated with aerobic exercise participation in stroke rehabilitation.

**Methods:** In this prospective cohort sub□study, adults admitted to in- or out-patient stroke rehabilitation at three urban hospitals completed validated questionnaires assessing depressive symptoms, apathy, exercise self□efficacy, outcome expectations for exercise, perceived barriers to being active, and premorbid exercise history. Participants were separated into two groups for analysis: those who completed aerobic exercise during rehabilitation and those who did not. Equivalence testing and between□group comparisons were performed.

**Results:** Sixty□two participants were enrolled; 16 participated in aerobic exercise and 46 did not. Groups were not equivalent on any individual□level factors. Compared to non□participants, those who performed aerobic exercise had significantly higher depressive symptom scores (p=0.0025) and lower self□efficacy for exercise (p=0.0087). Non□participants demonstrated significantly higher apathy (p=0.0007). No significant differences were found for outcome expectations, perceived barriers, or exercise history.

**Conclusion:** Depressive symptoms and lower self□efficacy did not impede aerobic exercise participation during rehabilitation. Increased apathy, however, was associated with non□participation. Findings highlight the need for individually tailored aerobic exercise prescriptions that consider motivational and affective factors to optimize engagement during stroke rehabilitation.

## INTRODUCTION

People with stroke often have low cardiorespiratory fitness which makes it challenging to complete some activities of daily living, such as carrying groceries and doing housework.^1^ Clinical guidelines recommend that aerobic exercise be incorporated into routine stroke rehabilitation to improve cardiorespiratory fitness.^2,3^ However, participation in structured aerobic exercise programs in stroke rehabilitation is low; only about a quarter of patients have aerobic exercise included in their treatment plans.^4^

Factors affecting participation in aerobic exercise during stroke rehabilitation are at the individual (patient), institutional (including therapist), or system levels; however, most studies have evaluated institutional or system level factors.^5–9^ Studies examining individual-level factors predicting physical activity or exercise participation in the community after discharge from stroke rehabilitation have found that self-efficacy for physical activity (i.e., confidence in one’s ability to perform physical activity), is one of the most reliable predictors.^10^ Lacroix et al., found that pre-stroke physical activity was significantly and positively correlated with physical activity during inpatient stroke rehabilitation.^11^ Patients reported that social supports, motivation to ‘get back to normal’, depressive symptoms, and fatigue influenced their ability to participate in stroke rehabilitation (which may or may not have included aerobic exercise).^12^ In contrast to these findings, in another study patients reported participating in any exercise that their physiotherapist prescribed to them (including aerobic exercise), regardless of how fatiguing or difficult the exercises were.^13^ The conflicting findings in this latter study mean that the evidence is unclear on whether the individual-level factors of self-efficacy for physical activity, exercise preferences, past exercise history and depressive symptoms influence participation in aerobic exercise during stroke rehabilitation.

People with stroke also have different exercise preferences than age-matched people who have not experienced a stroke. For example, people with stroke living in the community preferred exercise to be more structured, and the exercises to be demonstrated to them compared to age-matched non-stroke controls.^14^ People with sub-acute stroke (i.e., ∼3 weeks post-admission to rehabilitation) prefer low intensity, less frequent, shorter exercise sessions with more breaks;^15^ these preferences differ from clinical guidelines.^16^ Tailoring exercise delivery to participants’ preferences can influence affective responses to exercise,^17^ in turn influencing exercise behaviour.^18^ Therefore, improved understanding of exercise preferences of people with stroke could help tailor interventions to sustain participation in exercise.

We aimed to determine if selected individual-level factors (i.e., depressive symptoms, apathy, self-efficacy, outcome expectations for exercise, perceived barriers to exercise, and past exercise history) influence participation in aerobic exercise during stroke rehabilitation. From our previous qualitative study,^13^ we hypothesized that people with stroke who do and do not participate in aerobic exercise during rehabilitation are equivalent on these factors. Our secondary objective was to report on exercise preferences within our cohort.

## METHODS

### Participants

This is a sub-study of a larger prospective cohort study,^19^ which aimed to determine clinical predictors of participation in aerobic exercise during stroke rehabilitation. This larger study involved chart review of all patients admitted to four urban hospitals in Ontario, Canada over a 1-year period; three of the four sites were included in this study. All patients included in the chart review with sufficient cognitive, communication, and language ability to complete the questionnaires were invited to participate in this sub-study. Eligibility was determined via consultation with the patients’ inter-professional healthcare team. We aimed to recruit participants as early as possible after admission to rehabilitation (i.e., prior to starting any exercise) so that questionnaire responses would not be influenced by exercise participation. Participants provided written informed consent prior to participation, and the research ethics boards of the participating sites approved the study (University Health Network protocol number: 20-5695, and Sunnybrook Research Institute protocol number: 3605).

### Data collection

Participants underwent an in-person assessment with a research assistant. Participants completed a questionnaire via interview asking about their demographics, living situation, and employment; items in this questionnaire were adapted from the Canadian Longitudinal Study on Aging.^20^ Participants also completed the following questionnaires: the Medical Outcomes Study Social Support Survey (MOS),^21^ the Center for Epidemiologic Studies Depression scale (CES-D),^22^ the self-report version of the Apathy Evaluation Scale (AES),^23^ the Short Self-Efficacy for Exercise (SSEE) scale,^24^ the Short Outcome Expectation for Exercise scale (SOEE),^24^ the Barriers to Physical Activity After Stroke Scale (BPAS),^25^ the Schmidt retrospective physical activity scale,^26^ and the Stroke Exercise Preference Inventory (SEPI).^27^ The 20-item CES-D questionnaire has good test-retest and inter-rater reliability among people at various stages of stroke recovery (Pearson’s r=0.76, ICC=0.86).^28,29^ The self-report version of the AES has good test-retest reliability among a population with various neurologic conditions, including stroke (Pearson’s r=0.76).^23^ While apathy and depression can co-occur, they are often distinct constructs among people with stroke.^30^ The SSEE is a four-item questionnaire where participants rate their confidence exercising through pain and fatigue, and when alone and depressed. The Short Outcome Expectation for Exercise (SOEE) scale is a five-item questionnaire where participants rate their beliefs regarding the benefits of exercise. The SSEE and SOEE are reliable among individuals with chronic stroke (R^2^ for individual items 0.47-0.79).^24^ The BPAS is a 14-item questionnaire where participants are asked if they agree or disagree with statements about factors that might prevent them from being physically active (e.g., “I am in pain”). The BPAS has good test-retest reliability among people across various stages of stroke recovery (ICC=0.91).^25^ The Schmidt retrospective physical activity scale captures pre-morbid physical activity behaviour, by asking about physical activity, activities of daily life (i.e., for work, transportation and household) and exercise type, frequency and duration from 30-49 years and 50+ years of age. This scale shows good agreement with other questionnaires regarding physical activity.^26^ The SEPI (Part A) is a 13-item questionnaire that was specifically developed to assess preferences for exercise post-stroke. Participants rate each item (e.g., “I like to exercise alone”) on a scale from 0-100%.

Cohort descriptors (e.g., age, sex, type of stroke, more affected side) and whether patients were prescribed aerobic exercise were obtained from participants’ hospital charts. Aerobic exercise is structured, repetitive physical activity performed at a prescribed dose (i.e., frequency, intensity, and duration) with the goal of improving or maintaining cardiorespiratory fitness.^16^ In the study sites, aerobic exercise is prescribed by physiotherapists; while all patients should be prescribed aerobic exercise,^3^ the decision to prescribe aerobic exercise for a specific patient may depend upon patient goals, impairments, and co-morbid conditions.^5,19^ The Chedoke-McMaster Stroke Assessment (CMSA) leg and foot scores,^31^ Montreal Cognitive Assessment (MOCA),^32^ and National Institutes of Health Stroke Scale^33^ were also extracted from participants hospital charts; if these items were not available in the hospital chart they were assessed by the research assistant.

### Data analysis

All analyses were conducted using R Studio (version 2025.05.0+496, Posit Software, PBC, Boston, Massachusetts, USA). Participants were categorized into one of two groups based on their aerobic exercise participation during rehabilitation. Participants who participated in at least one session of aerobic exercise during rehabilitation were categorized into the group who participated in aerobic exercise, otherwise they were categorized into the group who did not participate in aerobic exercise. Continuous variables were tested for normality using Shapiro-Wilk’s test. Cohort descriptors were compared between groups using t-tests or Wilcoxon-Mann-Whitney tests (continuous variables) or Fisher’s exact test (categorical variables). Participant exercise preferences (SEPI Part A) were reported using descriptive statistics.

Total scores for the CES-D, AES, SSEE, SOEE, and BPAS were calculated. If participants reported any pre-morbid sport/exercise participation on the Schmidt retrospective physical activity questionnaire, regardless of level of exercise participation, they were considered as someone with exercise history. Exercise history was therefore treated as a binary variable.

Two one-sided tests (TOST) of equivalence and tests of difference, using either t-tests or Wilcoxon-Mann-Whitney tests (continuous variables) or chi-square test (exercise history) were conducted using the TOSTER package (version 0.8.4).^34,35^ The equivalence limits were: 3.8 for the CES-D (standard error of measurement),^29^ 3.74 for the AES (standard error of measurement), 0.5 for the SSEE and SOEE (e.g., 1 point difference in 2 questions), 4.47 for the BPAS (standard error of measurement),^25^ and 0.3 for exercise history (i.e., 30% difference in past exercise history between groups). Alpha was adjusted for multiple comparisons using the Holm-Bonferroni method,^36^ with the initial alpha set to 0.0083.

### Sample size calculations

Sample sizes were calculated using the TOST equivalence testing spreadsheet (version 0.4.6).^34,37^ For sample size calculations based on t-tests, we used the equivalence limits reported above, and standard deviations reported in previous publications.^23,25,29,38^ For exercise history, we assumed that 50% of participants would have a history of exercise participation. For all sample size calculations, alpha was 0.025 and beta was 0.8. Calculated sample sizes ranged from 31 per group (SOEE) to 234 per group (BPAS). Combined, the sites admit approximately 930 patients per year. From previous experience, we estimated that ∼50% of patients would be eligible for and consent to this data collection.^4,39,40^ Therefore, we expected a total sample size of ∼465.

## RESULTS

Recruiting and data collection occurred between 11 January 2021 and 27 November 2023 at the three sites. Of 981 people admitted to stroke rehabilitation at the sites during the recruiting time period, 19 were not included in larger study (e.g., because charts could not be retrieved) and 40 opted out of chart review for the larger study; these 59 patients were not considered for inclusion in the current study. From the remaining 922 patients, 62 were eligible and consented to participate in this study. Of these, 16 participated in aerobic exercise during rehabilitation and 46 did not. Cohort descriptors for the two groups are shown in Table 1.

**Table 1.** Participant characteristics extracted from patients’ charts. Values presented are means with standard deviations in parentheses (age), medians with interquartile ranges in parentheses (all other continuous variables), or counts with percentages in parentheses (categorical variables). The p-value is for the t-test comparing means between groups (age), Wilcoxon-Mann-Whitney test comparing medians between groups (all other continuous variables), or Fisher’s exact test comparing proportions between groups (categorical variables).

|  | Did aerobic exercise |  | Did not do aerobic exercise |  | p-value |
| --- | --- | --- | --- | --- | --- |
|  | N | Value | N | Value |  |
| Age (years) | 16 | 57.5 (16.6) | 46 | 62.0 (13.4) | 0.33 |
| Gender identity (n, %) | 16 |  | 46 |  | 0.37 |
| Female |  | 7 (43.8) |  | 14 (30.4) |  |
| Male |  | 9 (46.3) |  | 32 (69.6) |  |
| Marital status (n, %) | 16 |  | 46 |  | 0.0042 |
| Single |  | 6 (37.5) |  | 5 (10.9) |  |
| Married/common-law relationship |  | 9 (56.3) |  | 25 (54.4) |  |
| Widowed |  | 1 (6.3) |  | 1 (2.2) |  |
| Divorced/separated |  | 0 (0) |  | 15 (32.6) |  |
| MOS total (score) | 16 | 75.7 (41.4) | 45 | 80.3 (31.6) | 0.79 |
| Pre-morbid employment status | 16 |  | 45 |  | 0.81 |
| Employed full time ($\geq 30$ hours/week) | | 11 (68.8) | | 21 (46.7) | |
| Employed part time ($< 30$ hours/week) | | 1 (6.3) | | 15 (33.3) | |
| Retired |  | 4 (25.0) |  | 3 (6.7) |  |
| Unemployed |  | 0 (0) |  | 5 (11.1) |  |
| On disability leave |  | 0 (0) |  | 1 (2.2) |  |
| Pre-morbid household income (n, %) | 13 |  | 26 |  | 0.81 |
| \$20,000 - \$50,000 | | 3 (23.1) | | 6 (23.1) | |
| \$50,000 - \$100,000 | | 5 (38.5) | | 6 (23.1) | |
| \$100,000 - \$150,000 | | 2 (15.4) | | 6 (23.1) | |
| > \$150,000 | | 3 (23.1) | | 8 (30.8) | |
| Pre-morbid personal income (n, %) | 11 |  | 32 |  | 0.40 |
| <\$20,000 | | 0 (0) | | 6 (18.8) | |
| \$20,000 - \$50,000 | | 4 (36.4) | | 7 (21.9) | |
| \$50,000 - \$100,000 | | 5 (45.5) | | 10 (31.3) | |
| \$100,000 - \$150,000 | | 2 (18.2) | | 5 (15.6) | |
| > \$150,000 | | 0 (0) | | 4 (12.5) | |
| Highest level of education (n, %) | 16 |  | 45 |  | 0.20 |
| Did not complete high school |  | 0 (0) |  | 5 (11.1) |  |
| Graduated high school or equivalent |  | 3 (18.8) |  | 2 (4.4) |  |
| Trade certificate/diploma or apprenticeship |  | 1 (6.3) |  | 3 (6.7) |  |
| Non-university certificate/diploma |  | 2 (12.5) |  | 6 (13.3) |  |
| University certificate below bachelor's |  | 0 (0) |  | 5 (11.1) |  |
| Bachelor's degree |  | 6 (37.5) |  | 8 (17.8) |  |
| University degree or certificate above bachelor's |  | 4 (25.0) |  | 16 (35.6) |  |
| Service (n, %) | 16 |  | 46 |  | 0.065 |
| In-patient |  | 8 (50.0) |  | 11 (23.9) |  |
| Out-patient |  | 8 (50.0) |  | 35 (76.1) |  |
| Time post-stroke at admission (days) | 16 | 12.5 (48.0) | 46 | 42.0 (79.0) | 0.054 |
| Stroke type (n, %) | 16 |  | 46 |  |  |
| Ischemic |  | 10 (62.5) |  | 37 (80.4) | 0.033 |
| Lacune |  | 2 (12.5) |  | 0 (0) |  |
| Hemorrhagic |  | 3 (18.8) |  | 9 (19.6) |  |
| Both ischemic and hemorrhagic |  | 1 (6.3) |  | 0 (0) |  |
| More affected side (n, %) | 16 |  | 46 |  | 0.70 |
| Left |  | 8 (50.0) |  | 24 (52.2) |  |
| Right |  | 7 (43.8) |  | 14 (30.4) |  |
| Both |  | 1 (6.3) |  | 4 (8.7) |  |
| Neither |  | 0 (0) |  | 4 (8.7) |  |
| NIH-SS (score) | 4 | 8 (7) | 24 | 1 (3.5) | 0.050 |
| CMSA-leg (score) | 10 | 4.5 (3.0) | 28 | 5.0 (1.0) | 0.84 |
| CMSA-foot (score) | 10 | 4 (2) | 28 | 4 (2) | 0.95 |
| MOCA (score) | 11 | 23 (8) | 35 | 26 (4) | 0.28 |

MOS=Medical Outcomes Study Social Support Survey; NIH-SS=National Institutes of Health Stroke Scale; CMSA=Chedoke-McMaster Stroke Assessment; MOCA=Montreal Cognitive Assessment.

### Missing data

Data were missing when participants declined to complete all or part of an outcome assessment. Participants with missing data were excluded from analysis of those variables for which data were missing. The number of participants for whom data are available are included in the Tables.

### Comparison between groups

The BPAS was normally distributed and therefore was analysed using a t-test. All other continuous variables were not normally distributed and were analysed using Wilcoxon-Mann-Whitney tests. Comparison of outcomes between groups is show in Table 2. While approximately 56% of participants in both groups reported pre-morbid participation in sport or exercise, the groups were not statistically equivalent for exercise history using the Holm-Bonferroni adjusted alpha (Z=2.065, p=0.019). The two groups were not statistically equivalent on any other variables. Participants who did aerobic exercise scored higher on the CES-D (indicating more depressive symptoms) than participants who did not do aerobic exercise (Z=3.02, p=0.0025). Participants who did not do aerobic exercise scored higher on the AES (indicating increased apathy; Z=3.38, p=0.0007) and SSEE (indicating higher self-efficacy for exercise; Z=2.62, p=0.0087) ompared to participants who did aerobic exercise. SOEE scores were also higher (indicating greater outcome expectations for exercise) for those who did not do exercise compared to those that did, but this difference was not statistically equivalent using the Holm-Bonferroni adjusted alpha (Z=2.32, p=0.020). There were no statistically significant differences between the two groups for the BPAS (t_30.82_=0.90, p=0.38) or exercise history (p=0.49).

**Table 2:**
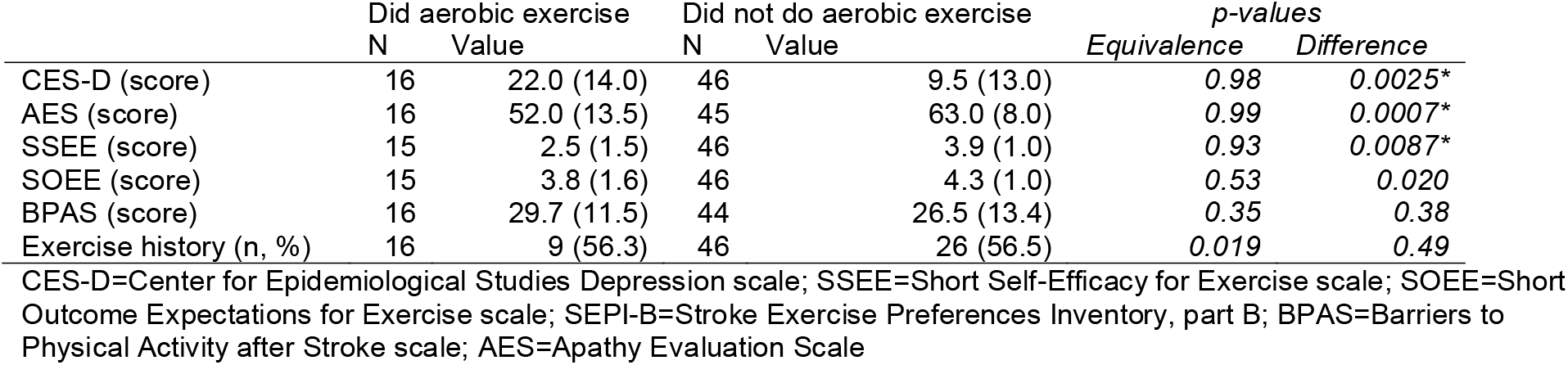
Results of equivalence and difference testing. Values presented are means with standard deviations in parentheses (BPAS), medians with interquartile ranges in parentheses (all other continuous variables) or number of people with a history of exercise with the percentage in parentheses (exercise history). P-values are for the two one-sided tests (TOST) test of equivalence and for the tests of differences. Statistically significant differences are indicated with an asterisk.

|  | Did aerobic exercise |  | Did not do aerobic exercise |  | *p-values* |  |
| --- | --- | --- | --- | --- | --- | --- |
|  | N | Value | N | Value | Equivalence | Difference |
| CES-D (score) | 16 | 22.0 (14.0) | 46 | 9.5 (13.0) | 0.98 | 0.0025* |
| AES (score) | 16 | 52.0 (13.5) | 45 | 63.0 (8.0) | 0.99 | 0.0007* |
| SSEE (score) | 15 | 2.5 (1.5) | 46 | 3.9 (1.0) | 0.93 | 0.0087* |
| SOEE (score) | 15 | 3.8 (1.6) | 46 | 4.3 (1.0) | 0.53 | 0.020 |
| BPAS (score) | 16 | 29.7 (11.5) | 44 | 26.5 (13.4) | 0.35 | 0.38 |
| Exercise history (n, %) | 16 | 9 (56.3) | 46 | 26 (56.5) | 0.019 | 0.49 |

CES-D=Center for Epidemiological Studies Depression scale; SSEE=Short Self-Efficacy for Exercise scale; SOEE=Short Outcome Expectations for Exercise scale; SEPI-B=Stroke Exercise Preferences Inventory, part B; BPAS=Barriers to Physical Activity after Stroke scale; AES=Apathy Evaluation Scale

### Exercise preferences

Participants’ exercise preferences (obtained from SEPI Part A) are listed in Table 3. The mean agreement for all the statements in the SEPI Part A was above 50%, except for liking to exercise with family or friends in the group that participated in aerobic exercise (49.4%), and liking to exercise with other people who have had a stroke in the group who did not participate in aerobic exercise (42.6%).

**Table 3:** Exercise preferences. Values are the mean % scores (standard deviations) for each item of the Stroke Exercise Preference Inventory Part A.

|  | Did aerobic exercise<br>(n=16) | Did not do aerobic<br>exercise (n=46) |
| --- | --- | --- |
| I like a trained instructor to supervise my exercise | 71.9 (43.2) | 62.9 (36.7) |
| I like to get feedback on how I'm going with my exercise | 67.5 (40.2) | 67.4 (36.9) |
| I am confident I can stay involved in a regular exercise program | 77.2 (36.6) | 76.3 (28.3) |
| I like to be challenged by exercises | 64.4 (38.3) | 73.0 (26.8) |
| I like to exercise for health reasons | 85.6 (27.1) | 90.1 (22.9) |
| It is important for me to do exercise that makes me feel good | 87.8 (27.0) | 88.5 (24.2) |
| I like to exercise with family or friends | 49.4 (41.8) | 52.9 (33.8) |
| I like to exercise outdoors | 54.8 (36.6) | 73.9 (33.5) |
| I like to exercise at home | 66.6 (36.5) | 71.4 (31.6) |
| I like to exercise alone | 66.3 (35.2) | 77.3 (29.2) |
| I like to exercise with other people who have had a stroke | 69.7 (39.5) | 42.6 (30.4) |
| I like to exercise with other people of similar age | 57.2 (41.6) | 51.4 (33.5) |
| I like to listen to music or watch TV during exercise | 79.7 (28.1) | 72.0 (36.3) |

## DISCUSSION

We investigated if people with stroke who participated in aerobic exercise during rehabilitation were equivalent to those who did not participate in aerobic exercise with regards to their depressive symptoms, apathy, exercise self-efficacy, outcome expectations for exercise, perceived barriers to exercise, and exercise history. Equivalence could not be declared for any of the outcome measures. Conversely, people who did not participate in aerobic exercise reported higher levels of apathy that those who did participate, and, somewhat unexpectedly, self-efficacy for exercise was lower and depressive scores were higher for those who participated in exercise compared to those who did not.

The findings of lower self-efficacy for exercise among those who participated in aerobic exercise conflicts with other studies reporting that self-efficacy for exercise^10,24,41–43^ and outcome expectations for exercise^41^ predict participation in self-directed community-based exercise post-stroke. Self-efficacy and outcome expectations are important influences on self-motivation for behaviour.^44^ However, participants in our study were attending stroke rehabilitation, with aerobic exercise provided by physiotherapists. To our knowledge, this is the first study comparing exercise self-efficacy in people with stroke who participate in aerobic exercise during rehabilitation with those who do not. Self-motivation to initiate exercise is less important in this setting than when people are living independently in the community. Within a rehabilitation context, patients seldom report low motivation to participate in exercises provided by their therapists.^9,13^ Higher self-efficacy among those who did not participate in aerobic exercise may suggest that these patients were physically fit or active outside of physiotherapy sessions, limiting physiotherapists’ perceived need to include exercise in their treatment.^5^ However, we did not have access to cardiorespiratory fitness or physical activity data outside of therapy sessions to explore this possibility. Furthermore, even if those with high exercise self-efficacy were physically active outside of therapy sessions, it is possible that the volume or intensity of this activity is not sufficient to improve cardiorespiratory fitness.^45^ Therefore, these patients with high exercise self-efficacy would likely still benefit from prescribed aerobic exercise during stroke rehabilitation.

Our findings of more depressive symptoms among those who were prescribed aerobic exercise conflict with others. Previous studies found that increased depressive symptoms were associated with reduced participation in stroke rehabilitation,^46^ and physical activity in the community.^47^ Our larger study, using the same population as the current study, found that people with diagnosed mental health conditions were less likely to complete aerobic exercise than those without mental health diagnoses.^19^ This discrepancy may be explained, in part, by the fact that high scores on the CES-D scale do not necessarily translate to diagnosis of depression,^48^ and that all mental health diagnoses (e.g., anxiety) were collapsed into a single variable in our larger study.^19^ Nonetheless, the results of the current study suggest that, while low mood may make exercise more difficult,^13^ depressive symptoms are not a barrier to participating in aerobic exercise during stroke rehabilitation.

Our unexpected findings regarding self-efficacy and depression could also be explained by the difference in time post-stoke between groups. There were more outpatients in the group that did not participate in aerobic exercise compared to the group that did; consequently, time post-stroke on admission was, on average, 12.5 days for the group who participated in aerobic exercise compared to 42 days for those who did not. There are significant improvements in body functions and activities in the first 6-10 weeks post stroke.^49^ Likewise, depression improves with recovery from stroke.^50^ Because people with stroke who did not participate in aerobic exercise in our study were further along their recovery path, they presumably had more improvements in terms of their motor function, activities of daily living and emotional and behavioural changes compared to those who did not participate in aerobic exercise.^49,50^

Our findings of increased apathy among those who did not participate in aerobic exercise agree with other studies showing that reduced apathy predicts participation in physical activity in the community post-stroke.^47^ Apathy is characterized by low motivation,^51^ which is a significant predictor of behaviour change.^52^ Participants with increased apathy may be less motivated to engage in aerobic exercise, even when it is prescribed by their therapists. However, our previous data from these sites suggest that patients rarely declined aerobic exercise when prescribed.^5,13^ Our previous work also found that physiotherapists view aerobic exercise as an ‘add on’ to therapies focused on physical function (e.g., transfers, ambulation).^5^ Patients with high apathy may be less engaged in rehabilitation in general, leading to physiotherapists being less likely to initiate ‘supplementary’ therapies, like aerobic exercise. Conversely, participation in exercise may improve apathy,^53^ so strategies to engage those with low motivation in exercise are needed.

In terms of exercise preferences, participants in our study generally did not like to exercise with others. In our previous qualitative study, one participant reported feeling uncomfortable exercising with others with stroke, due to the tendency to compare progress with other patients.^13^ Conversely, others have reported that social support from other patients, and seeing others progress can facilitate them participating in stroke rehabilitation.^12^ Most of the participants in our study (>70%) were interested in listening to music or watching television during exercise. Bonner et al. found that participants with more depressive symptoms have a greater preference for music or television during exercise.^27^ Additionally, a recent meta analytic review reported that listening to music is associated with more positive feelings, improved physical performance, reduced perceived exertion, and more efficient use of oxygen.^54^ The confidence-challenge factor (i.e., “I like to be challenged by exercises”, and “I am confident I can stay involved in a regular exercise program”) is closely related to exercise self-efficacy.^55^ Interestingly, the group who participated in aerobic exercise in our study had a slightly lower tendency to be challenged by exercise compared to the group who did not (64.4 % agreement with the statement vs 73%), which agrees with the lower exercise self-efficacy score seen in this group.

This study is limited by a smaller sample size than required for 80% power, thus, the results need to be considered with caution. Our sample size was low due to lower than expected rates of participation in the study. Underpowered tests increase the risk of inconclusive results in equivalence studies more than traditional hypothesis tests where large effects can compensate for small sample sizes.^56^

## Conclusion

This study investigated whether selected individual-level factors (i.e., self-efficacy, perceived barriers to exercise, apathy, past exercise history and depressive symptoms) influence participation in aerobic exercise during stroke rehabilitation. While we did not reach recruitment targets, we did not find any evidence that low self-efficacy or depressive symptoms are significant barriers to participation in aerobic exercise during in-patient rehabilitation. However, between-group differences in apathy suggest that this factor may prevent aerobic exercise participation.

## Data Availability

Data are not available publicly due to local privacy legislation.

## Notes

**Funding:** This study was supported by the Canadian Institutes of Health Research (PJT 173472).

### Competing Interest Statement

The authors have declared no competing interest.

### Author Declarations

The research ethics board of the University Health Network (protocol number: 20-5695) and the Sunnybrook Research Institute (protocl number: 3605) gave ethical approval for this work

